# Superspreading in Early Transmissions of COVID-19 in Indonesia

**DOI:** 10.1101/2020.06.28.20142133

**Authors:** Agus Hasan, Hadi Susanto, Muhammad Firmansyah Kasim, Nuning Nuraini, Bony Lestari, Dessy Triany, Widyastuti Widyastuti

## Abstract

We estimate the basic reproduction number *ℛ*_0_ and the overdispersion parameter *𝒦* at two regions in Indonesia: Jakarta-Depok and Batam. Based on the first 1288 confirmed cases in both regions, we find a high degree of individual-level variation in the transmission. The basic reproduction number *ℛ*_0_ is estimated at 6.79 and 2.47, while the overdispersion parameter *𝒦* of a negative-binomial distribution is estimated at 0.06 and 0.2 for Jakarta-Depok and Batam, respectively. This suggests that superspreading events played a key role in the early stage of the outbreak, i.e., a small number of infected individuals are responsible for large amounts of COVID-19 transmission.

## Introduction

The first two confirmed cases of COVID-19 in Indonesia was announced in March 2, 2020 by the president himself. Jakarta and its buffer zones including Depok have become the epicenter of the outbreak. In its early transmission, about 50% of the cases were from the cities^1^. The virus quickly spread to all 34 provinces, with more than 93,000 confirmed cases and 4,500 deaths as of July 23, 2020. As a country with the highest death toll in South East Asia and one with the lowest global testing rate, the first government effort to control the disease spread was by establishing a COVID-19 response acceleration task force led by the head of the National Disaster Management Agency. The government further introduced Large-Scale Social Restrictions, which included measures such as closing public places, restricting public transport, and limiting travel to and from restricted regions. On May 19, the first milestone of 10,000 PCR tests per day was reached. In June 2020, WHO announced that only Jakarta meets the minimum requirement of 1 test per 1,000 population per week for a reliable positivity rate calculation^2^.

As the Indonesian government still struggles to mitigate the COVID-19 epidemic in the country, it becomes critical to understand its infection spreads. Here, we present a first quantitative analysis of early transmission dynamics of the disease and evidence of superspreading events in Indonesia based on contact tracing of secondary cases. Infection clusters are difficult to predict and thus difficult to prevent. Nevertheless, understanding their environmental and behavioral drivers has been recently recognized to be extremely important to inform strategies for prevention and control^3^ as the reproduction number only is no longer enough^4^. In our analysis of Jakarta, we combine data from Depok that is a main buffer city of the capital. Outside the epicentre, we present data from Batam that has received thousand test kits and protective equipment from Singapore as a comparison. There were 218 cases from 4,092 PCR tests, as of June 26^5^.

## Results

Based on 1288 confirmed cases in March and April 2020 in the regions Jakarta-Depok^1^ (population 12.6 millions) and Batam^5^ (population 1.4 millions), we find a high degree of individual-level variation in the transmission. Early in the outbreak, superspreaders transmit the majority of coronavirus cases, as can be seen from Figure 1. Defining a superspreader as an individual with at least eight transmissions of the disease, there were 44 clusters in Jakarta-Depok region (2 - 31 March, 2020) and 3 clusters in Batam city (19 March to 7 April, 2020), as can be seen from Table 1.

**Table 1.** Secondary cases data of the two regions based on local transmissions.

| Jakarta-Depok |  | Batam |  |
| --- | --- | --- | --- |
| Secondary cases | Frequency | Secondary cases | Frequency |
| 0 | 1017 | 0 | 63 |
| 1 | 31 | 1 | 12 |
| 2 | 36 | 2 | 3 |
| 3,6 | 17 | 3 | 4 |
| 4,13 | 4 | 4,7-9,13 | 1 |
| 5 | 22 | 5 | 2 |
| 7 | 11 |  |  |
| 8,11,12 | 3 |  |  |
| 9 | 8 |  |  |
| 10 | 7 |  |  |
| 14,17,20,21,26,27,36,43 | 1 |  |  |
| 15,16,19,23 | 2 |  |  |
| Total | 1199 | Total | 89 |

**Figure 1.**
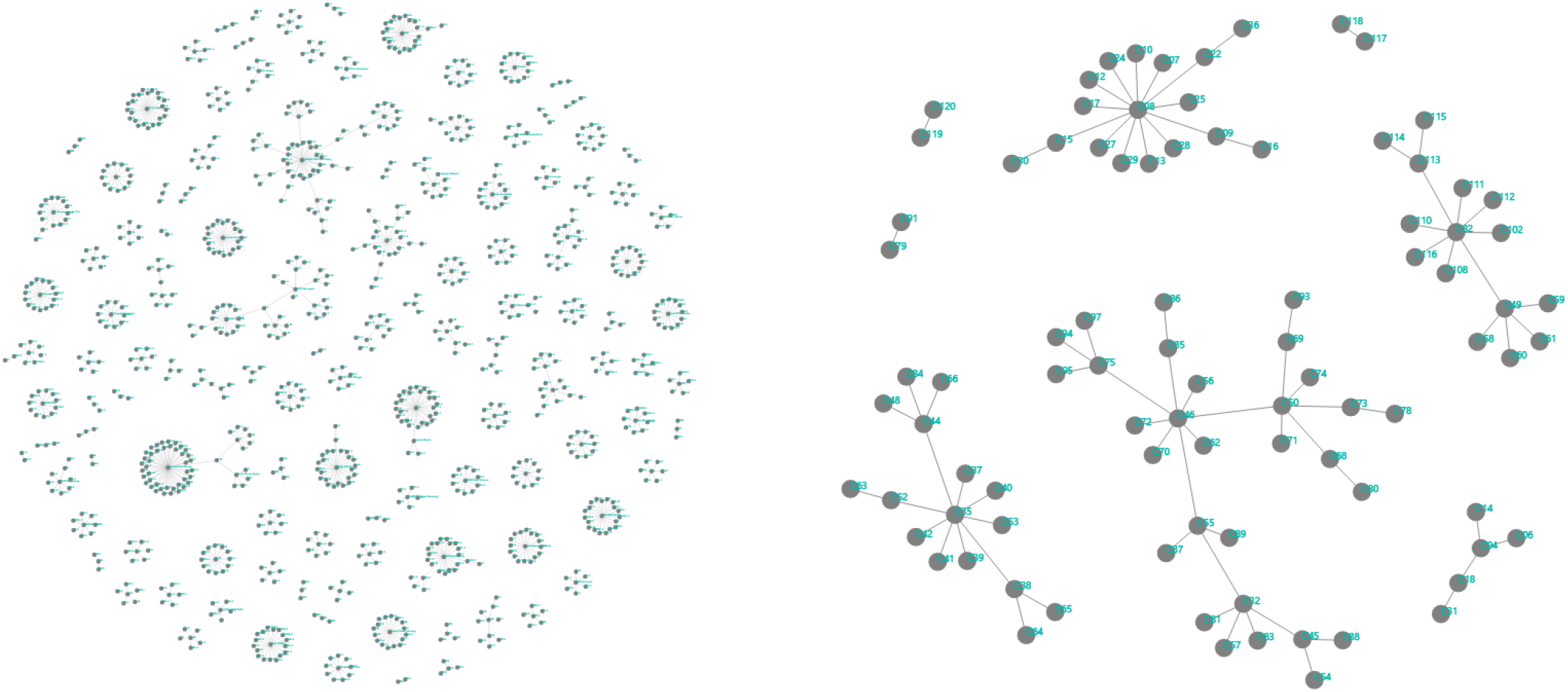
Networks of secondary cases based on local transmissions in Jakarta-Depok (left) and Batam (right) regions.

We estimated the reproduction number *ℛ*_0_ to be at 6.79 and 2.47, while the overdispersion parameter *𝒦* of a negative-binomial distribution is obtained at 0.06 and 0.2 for Jakarta-Depok and Batam, respectively (see Figure 2), suggesting that a small number of infected individuals are responsible for large amounts of the disease transmission. Indeed, between 10-15% of all infections were responsible for 80% of onward transmission events.

**Figure 2.**
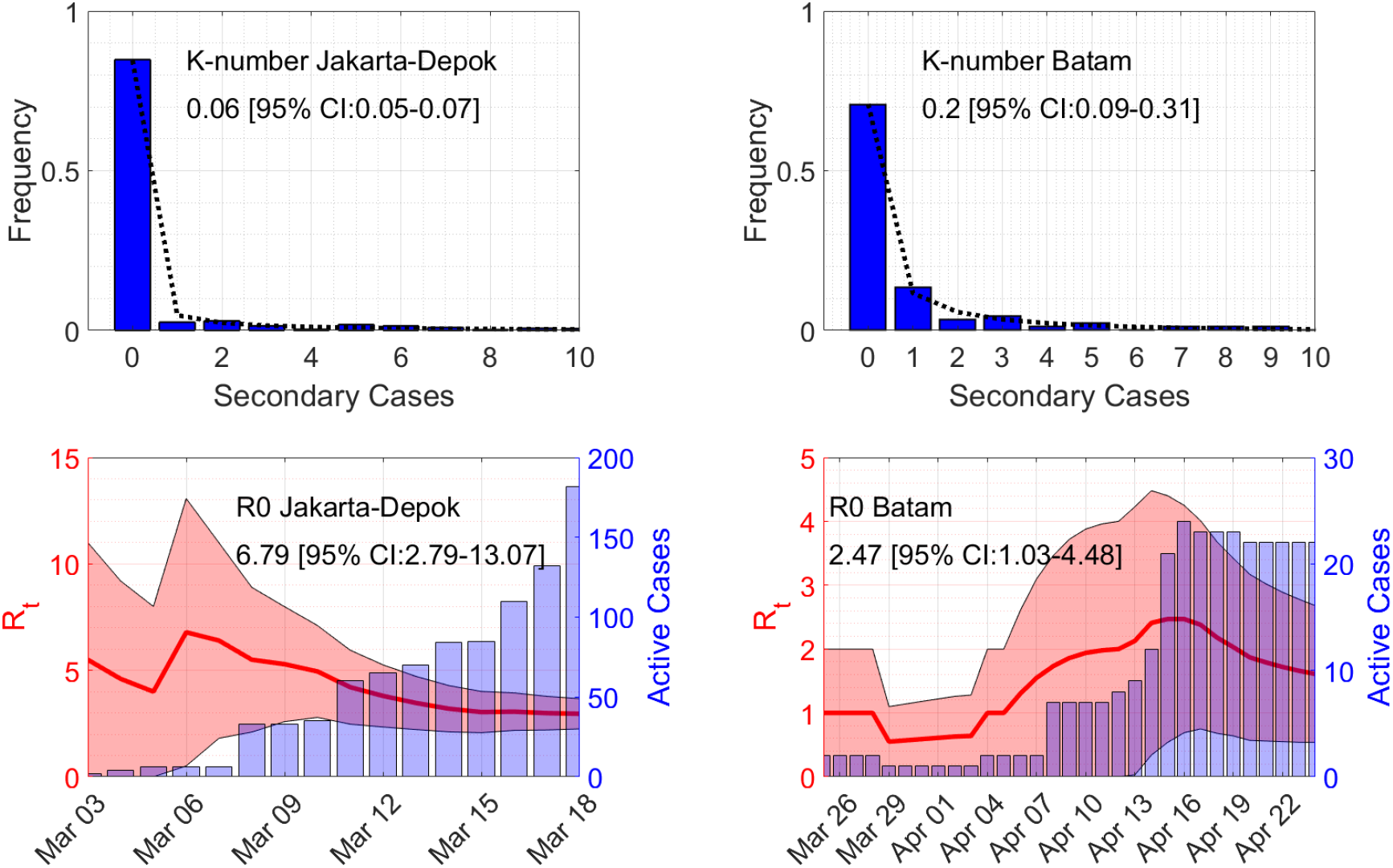
Estimates of the overdispersion parameter *𝒦* and the basic reproduction number *ℛ*_0_. Lines in the top figures show maximum-likelihood fits for the negative binomial distribution.

The fact that Indonesia has a moderate number of positive cases, compared to other countries with roughly the same population size such as Russia, USA, and Brazil, can be caused by three reasons: a low number of tests, the success of Large-Scale Social Restrictions, and overdispersion of the COVID-19 transmission. Our result clearly indicates that the transmission is overdispersed, even though it does not exclude the other possibilities. Therefore, close-interaction activities such as traditional markets, religious gathering, and wedding parties need to be adapted if not restricted as they can become transmission hot spots. Effective response factors to reduce the transmission include aggressive implementation of non-pharmaceutical interventions such as rapid identification and isolation of cases. Furthermore, timeliness is critical to prevent or limit their extent since delay of diagnosis is the most common cause of superspreading clusters^6^.

## Methods

We use a sequential Bayesian method based on the discrete SIR model to estimate the basic reproduction number *ℛ*_0_^7^. Time series of the active cases at day *n* is assumed to follow the model *I*_*n*+1_ = (*γ* (ℛ_*t*_ *−* 1) + 1) *I*_*n*_, where *ℛ*_*t*_ is the instantaneous reproduction number and 1*/γ* is the infectious time taken to be 9 days herein^8^. Assuming a Poisson distribution for the arrival of active cases and using Bayes’ theorem, the posterior probability of *ℛ*_*t*_ given a confirmed case *I*_*n*+1_ is computed through

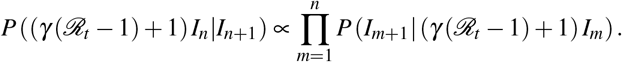

The formula comes from the assumption that the prior distribution of *ℛ*_*t*_ at day *n* is taken from the posterior of day (*n −* 1) with a uniform prior at day 1. Estimated *ℛ*_0_ is then the maximum of *ℛ*_*t*_, see Figure 2.

The parameter *𝒦* is then defined as the overdispersion parameter in the negative binomial distribution^9^, which can be found by fitting the distribution to the secondary cases data. The number of secondary cases by one person *x* can be expressed in terms of the overdispersion parameter *𝒦* and the probability *p*,

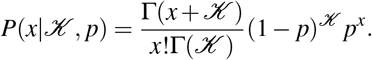

We use a MATLAB^®^ function *nbinfit* to estimate the overdispersion parameter *𝒦*. The function uses derivative-free method to find the maximum of the likelihood function.

## Data Availability

Data are available in the manuscript.

## Data and Code Availability

Secondary cases data (both in Excel and Power BI) and a MATLAB^®^ code to produce Figure 1 and Figure 2 are available at: https://github.com/agusisma/covidindonesia.

## Acknowledgements

We would like to thank Public Health Office of Jakarta Province, Communication, Information and Statistic Office of Jakarta Province, Jakarta Smart City Unit, Human Settlements and Spatial Planning Office of Jakarta Province, Public Health Offices of Batam, and Depok Task Force for COVID-19 Control for providing the data, feedback, and discussions.

## Author contributions statement

A.H., H.S., and M.F.K. analysed the data, calculated *ℛ*_0_ and *𝒦*, and discussed the results, N.N., B.L., D.T., and W.W. collected the data. All authors reviewed the manuscript.

